# Re-evaluating the evidence for fecal microbiota transplantation “super-donors” in inflammatory bowel disease

**DOI:** 10.1101/19011635

**Authors:** Scott W. Olesen, Ylaine Gerardin

## Abstract

**Background:** Fecal microbiota transplantation (FMT) is a recommended treatment for recurrent *Clostridioides difficile* infection, and there is promise that FMT may be effective for conditions like inflammatory bowel disease (IBD). Previous FMT clinical trials have considered the possibility of a “donor effect”, that is, that FMT material from different donors has different clinical efficacies.

**Aim & Methods:** Here we re-evaluate evidence for donor effects in published FMT clinical trials for IBD.

**Results:** In 7 of 9 published studies, no statistically significant donor effect was detected when rigorously re-evaluating the original analyses. One study had statistically significant separation of microbiota composition of pools of donor stool when stratified by patient outcome. One study reported a significant effect but did not have underlying data available for re-evaluation. When quantifying the uncertainty on the magnitude of the donor effect, confidence intervals were large, including both zero donor effect and very substantial donor effects.

**Conclusion:** Although we found very little evidence for donor effects, the existing data also cannot rule out the possibility that donor effects are clinically important. Large clinical trials prospectively designed to detect donor effects are likely necessary to determine if donor effects are clinically relevant for IBD.

## INTRODUCTION

The human microbiome is increasingly understood to play a key role in health and disease.^1^ Fecal microbiota transplantation (FMT), the infusion of a healthy person’s stool into a patient, is one method for manipulating the gut microbiome.^2^ FMT is recommended for treatment of recurrent *Clostridioides difficile* infection.^3,4^ Although our understanding of the specific mechanisms by which FMT cures *C. difficile* infection are still developing, FMT is being investigated as a therapy for dozens of other microbiome-related indications.^2,5,6^

A key challenge in identifying FMT’s specific mechanism, or mechanisms, is the complexity and diversity of human stool. Stool is a mixture of bacteria, viruses, fungi, microbe-derived molecules, and host-derived molecules that varies enormously from person to person.^7,8^ It has therefore been hypothesized that different stool donors, different FMT material, or different matches of donors and recipients could have different abilities to treat disease. This concept has been referred to with terms like “donor effect”, “super-donor”, and “super-stool”.^6,9–12^

If FMT’s efficacy varied widely across stools or donors, then rational selection of FMT material based on biomarkers predictive of efficacy could improve the clinical practice of FMT.^6,11,13,14^ Differences in FMT efficacy between stool donors or specific stools could be also an important starting point for scientific investigations into the “active ingredient” in FMT donations.^11^ It is therefore important to quantify the extent of donor variability in indications where FMT is a promising treatment.

Although there is little evidence of a donor effect in the context of *C. difficile* infection,^15–17^ the condition for which the use of FMT is best studied, multiple studies using FMT to treat inflammatory disease have tested for donor effects,^2,9,10,18–24^ and some have reported statistically significant results. However, there are reasons to be skeptical of the clinical implications of these intriguing findings.

First, a previous report^25^ found that, in simulations of clinical trials, studies with numbers of patients similar to the number used in extant studies would be unlikely to discover a donor effect unless it were very large. Statistically significant results from underpowered studies may represent discoveries of real effects, but they may also represent false positives.

Second, in all previous studies, identifying a donor effect was a *post hoc* analysis, and in no case was multiple hypothesis correction, a critical methodology in *post hoc* analyses, employed. In addition, we show herein that, in at least two previous reports, the specifics of the clinical design require adjustments to the statistical tests used to avoid inflating the apparent evidence for a donor effect.

Third, previous reports mostly analyzed donor effects by testing null hypotheses that are biologically unlikely: either that all donors produce stool with exactly equal clinical efficacy, or that particular features of stool, like its bacterial community diversity, are completely unrelated to clinical efficacy. However, it is highly improbable that all stool has precisely the same therapeutic effect: if a study is large enough, the difference between two donors’ treatment efficacies would almost certainly become statistically significant as the number of treated patients increases. We therefore propose that, rather than merely asking if donor effects exist or not, we should ask about the magnitude of the donor effect and whether it is clinically relevant. In other words, rather than merely testing for the statistical significance of donor effects, we should also assess effect sizes.^26^

Here we re-evaluate the existing evidence for donor effects and discuss the implications of that evidence for clinical trial design and clinical practice. First, we establish an ontology of the various concepts referred to as “donor effects”. Second, we lay out a rigorous framework for identifying and quantifying donor effects. Third, we re-evaluate the existing literature using the conceptual ontology and the rigorous framework. Finally, we discuss the implications of this re-evaluation for future FMT research in IBD.

## METHODS

### Distinguishing types of stool superiority

Discussions of “super-donors” and “super-stool” have suggested that stool might be superior in at least 4 distinct but conceptually related senses. To avoid confusion in our re-evaluation of the evidence for donor effects, we distinguish between these definitions of stool superiority:

1. *Donor superiority*, in which particular donors are associated with better clinical outcomes for the recipient patients. For example, Moayyedi *et al*.^18^ tested whether a particular donor (“donor B”) was associated with better patient outcomes, compared to the other donors.
2. *Donor characteristic superiority*, in which donors with particular characteristics are associated with better outcomes. For example, donor age, diet, and host genetics have been suggested as potential factors in donor superiority.^10,24,27^
3. *Material characteristic superiority*, in which stool donations, pools of stool, or other fecal microbiota preparations that have some particular characteristic are associated with better outcomes. For example, Vermeire *et al*.^28^ tested whether stools with higher bacterial diversity are associated with better outcomes.
4. *Donor-recipient match superiority*, in which certain combinations of donors and recipients are associated with better outcomes,^9,10,21,29^ analogous to how donors’ and recipients’ blood types are matched for blood transfusions. For example, FMT studies have tested whether stool from “related” donors,^30^ typically defined as first-degree relatives but also sometimes including spouses or partners, is associated with better or worse patient outcomes.

These types of superiority are conceptually related and not mutually exclusive. For example, imagine it were the case that female donors were associated with better patient outcomes than male donors, consistent with definition 2, “donor characteristic superiority”. However, because donors have a clinical effect on the patient only via their donation, sex of donor cannot be the molecular mechanism by which some donations are more efficacious than others. It would have to be that sex determined or correlated with some component of the stool that made those donations more effective. In other words, there must be an underlying “material characteristic superiority”, definition 3, that correlates with donor sex. By a similar argument, even if donor-recipient match superiority is the most accurate model, simpler types of superiority may be more parsimonious.^12^

### Identifying and quantifying donor effects

We used a two-step framework for characterizing donor effects. First, we used statistical tests to characterize the strength of the evidence for the reported type of donor effect. Statistics were computed using the R programming language^31^ (version 3.6.0). To test the hypothesis that there is any difference in efficacy between donors (or pools of donors), we used the Fisher-Freeman-Halton test (function *fisher.test*) on 2 × *D* contingency tables, where *D* is the number of donors (or pools). For 2 × 2 contingency tables (e.g., to test if one donor has a different treatment efficacy than all the others), we used Fisher’s exact test (function *exact2*×*2*)^32^ and the mid-p value to prevent the tests from being overly conservative.^33^ To test for material (i.e., donation or pool) characteristic superiority, we used the Mann-Whitney *U* test (*wilcox.test*) or Kruskal-Wallis test (*kruskal.test*). When 16S rRNA gut microbiota composition data were available (see below), we tested for separation in donor microbiota compositions using PERMANOVA^34^ (function *Adonis* in the R *vegan* package).^35^

Second, whether or not an effect was detected, we use a random effect logistic regression (function *glmer* in the R *lme4* package)^36^ to quantify the effect size. The regression model assumes that the log odds of the efficacy of different donors (or pools of stool) is Gaussian distributed. The regression returns the standard deviation of that distribution, which is a measure of the “spread” in donors’ efficacies. For clarity, we convert the regression’s estimates into the absolute difference in efficacy rates between a 90th percentile donor (a very high quality donor) and a 10th percentile donor (a poor donor). For example, an effect size of 30 percentage points means that, if a low quality donor is 10% efficacious, then a high quality donor would be 40% efficacious.

### Studies re-analyzed

In the re-analysis, we included interventional FMT clinical studies that tested the hypothesis that one donor or donation characteristic was associated with improved IBD patient outcomes. Studies were identified using citations from an existing systematic review,^21^ narrative review,^10^ and expert opinion.^6^ For each study, we considered only a single dichotomous outcome, whether a patient achieved the trial’s primary clinical endpoint. When 16S rRNA gut microbiota composition data were available, we tested for associations between donor material α-diversity and patient outcomes,^23,24,28^ since part of the motivation for using pools of stool from multiple donors is the hypothesis that high bacterial diversity in FMT material is beneficial.^13,19,29,37^ When possible, we also searched for separation in the microbiota compositions of donors according to the outcomes of their associated patients, following the example of Jacob *et al*.^20^

### Microbiome data preparation

We re-analyzed raw 16S rRNA sequencing data from three studies (Kump *et al*., study accession PRJEB11841; Jacob *et al*., accession PRJNA388210; Goyal *et al*., accession PRJNA380944) using QIIME 2^38^ (version 2019.7) and Deblur.^39^ For Kump *et al*. and Jacob *et al*., paired reads were joined (*vsearch* plugin, default parameters), quality filtered (*quality-filter* plugin, default parameters), and denoised using Deblur (trim length 253 nucleotides, 1 minimum reads, otherwise default parameters). For Goyal *et al*., the sequencing data was single-ended (no joining required) and the trim length was 150 nucleotides. Alpha-diversity was computed by down-sampling all samples to the minimum number of denoised read counts across samples and then using the Shannon metric (*diversity* plugin). When a donor had multiple associated samples, we associated a single α-diversity with each donor by computing the mean α-diversity over that donor’s samples. Beta-diversity, used for the PERMANOVA tests, was computed with the Bray-Curtis metric. When a donor had multiple samples, we used only the first sample from each donor.

### Code and data availability

Computer code and underlying data to reproduce the results are online (DOI: 10.5281/zenodo.3780184, https://www.github.com/openbiome/donor-effects).

## RESULTS

The results of the re-evaluation of the evidence for a donor effect in IBD from a selection of FMT clinical studies in summarized in Table 1.

**Table 1:** Summary of re-analyses.

| Data source | No. patients | No. donors | Evidence for donor superiority?<sup>a</sup> | Evidence for pool superiority?<sup>a</sup> | Association between donor α-diversity and outcomes?<sup>b</sup> | Separation in donor microbiota composition by patient outcome?<sup>c</sup> |
| --- | --- | --- | --- | --- | --- | --- |
| Rossen *et al.* | 23 | 15 | No | — | — | — |
| Moayyedi *et al.* | 38 | 6 | No | — | — | — |
| Paramsothy *et al.* | 78 | 14 | No | No (0 p.p., 95% CI 0 to 46) | — | — |
| Costello *et al.* | 38 | 19 | No | No (0 p.p., 95% CI 0 to 74) | — | — |
| Jacob *et al.* | 20 | 4 | Yes ($p = 0.02$) | No (88 p.p., 95% CI 0 to 100) | No | No |
| Pool meta-analysis<sup>‡</sup> | 137 | 36 | No | No (41 p.p., 95% CI 0 to 88) | — | — |
| Goyal *et al.* | 21 | 21 | — | — | No | No |
| Kump *et al.* | 17 | 14 | — | — | No | No |
| Vermeire *et al.* | 14 | 14 | — | — | Reported | — |
| Nishida *et al.* | 41 | 41 | — | — | No | — |

p.p.: Percentage points
a: Tests of donor superiority and pool superiority were using Fisher-Freeman-Halton or Fisher's exact test. Quantifications of pool effect are differences in clinical efficacy between a very high quality pool (90th percentile) and low quality pool (10th percentile).
b: Using Mann-Whitney *U* test or Kruskal-Wallis test
c: Using PERMANOVA on Bray-Curtis β-diversity

### Rossen *et al*. 2015

In this study,^9^ 23 ulcerative colitis patients were treated with FMT. 17 patients received stool from 1 of 15 donors; 6 patients received stool from 2 different donors. The outcome was clinical remission and endoscopic response at 12 weeks. In the original publication, the investigators reported on the performance of 3 donors (Table 2). There was no evidence of a donor effect (mid-*p* > 0.05, Fisher’s exact test) for any of the 3 donors. However, the study is small enough that the data are not inconsistent with large variability between donors. For example, the upper 95% confidence interval on donor A’s odds ratio of successful patient outcome compared to all other donors is 29. In other words, the data do not provide evidence of a donor effect, but they also cannot definitively rule out a strong donor effect.

**Table 2:** Tests of donor superiority, by donor, for Rossen *et al*. Donor labels are arbitrary. “Success” means the patient reached the primary endpoint; “failure” means they did not.

|  | Patients treated using this donor |  | Patients treated not using this donor |  | Fisher’s exact test |  |
| --- | --- | --- | --- | --- | --- | --- |
| Donor | Success | Failure | Success | Failure | Odds ratio (95% CI) | Mid-*p* value |
| A | 4 | 4 | 3 | 12 | 3.7 (0.55 to 29) | 0.18 |
| B | 1 | 2 | 6 | 14 | 1.2 (0.03 to 18) | 0.89 |
| C | 0 | 2 | 7 | 14 | 0.0 (0.0 to 8) | 0.47 |

### Moayyedi *et al*. 2015

In this study,^18^ 38 ulcerative colitis patients were treated with FMT. Each patient was treated with stool from 1 of 6 donors. The outcome was remission at 7 weeks. Donors were used according to an adaptive process, described in the original publication. Briefly, at the start of the trial, patients were treated with stool from one of two donors (A or B). Material from donor B then became unavailable, and patients were instead treated with stool from donor A or 1 of 4 other donors. At this point, donor B became available again, and “[t]he remaining participants allocated to active therapy all received FMT from donor B exclusively, as [the investigators] had not experienced any success with donor A”. In the original study, donor B’s performance (7 of 18 treated patients achieved the primary endpoint) was compared against all other donors (2 of 20 patients; odds ratio 5.5 [95% CI 1.0 to 44], mid-*p* = 0.048, Fisher’s exact test).

In general, adaptive trial designs require special statistical methodologies.^40^ To investigate whether typical statistical tests like Fisher’s exact test accurately measure the statistical significance of the data collected in Moayyedi *et al*., we simulated a simplified adaptive donor selection process. First, among the first 24 simulate patients, use all 6 donors 4 times each. Then select the best-performing donor to treat all the remaining 14 patients. Finally, compare the overall performance of this donor against all the others. In these simulations, if all the donors are actually the same (i.e., with equal probability 9/38 = 24% of a positive patient outcome), the probability of finding mid-*p* < 0.05 is 8.8% (8843 of 10,000 simulations; 95% CI 8.7% to 9.0%), nearly double the value that would be expected if the test were accurate for that type of data (i.e., the false positive rate, 5%). The intuitive explanation is that, if all donors are the same, the selection process is merely setting aside donors who had “bad luck” on their first patients, and they are not allowed to recover their performance with a run of “good luck” later on. This is not a flaw of Fisher’s exact test. Instead, it is problem of applying a statistical test to data that do not meet the assumptions of the test.

Although the adaptive procedure that led to extensive use of donor B may have improved the probability of the trial’s success,^12^ the procedure was not pre-specified, making it impossible to rigorously determine the degree to which the observed results are consistent with all donors being identical, that is, whether donor B simply had a “lucky” initial run. Thus, the value mid-*p* = 0.05 reported above is inaccurately optimistic, and a *p*-value of 0.09 is a better representation of the likelihood of a donor having the kind of success observed in this trial.

### Paramsothy *et al*. 2017

In this study,^19^ 41 ulcerative colitis patients were randomized to receive blinded FMT. Another 37 were randomized to placebo and later received open-label FMT. Each of the 78 patients received FMT from 1 of 21 pools of donors. Each pool included material from 3 to 7 donors, out of 14 total donors. The outcome in this analysis was clinical remission with endoscopic remission or response at 8 weeks after either the blinded or open-label FMT.

The original publication compared the best performing individual donor against all the others (14 of 38 patients achieved the primary outcome, versus 7 of 40 assigned to other donors; odds ratio 2.7 [95% CI 0.96 to 8.2], mid-*p* = 0.06, Fisher’s exact test). This result, although near mid-*p* = 0.05, is not convincing when subjected to multiple hypothesis correction, as there are 14 relevant hypotheses to be tested, one per donor (Bonferroni-corrected *p* = 1.0, Benjamini-Hochberg FDR = 0.78). In terms of pools of donors, there was also no evidence that any particular pool was associated with better outcomes (*p* = 0.76, Fisher-Freeman-Halton test on 2 × 21 table of patient outcomes by pool).

To develop an estimate of the strength of the donor effect, we used a logistic regression, modeling the pool as a random effect. This model estimated that the difference in efficacy between a very high quality pool (90th percentile in terms of efficacy) and a poor pool (10th percentile) is exactly 0 percentage points (95% CI 0 to 46 percentage points). In other words, the regression’s best estimate of the variation in efficacy among pools is zero, even without applying any penalizations for model complexity. Thus, similar to Rossen *et al*., the data do not provide evidence for a donor effect, but they are also not inconsistent with a strong effect, since the upper confidence limit on the strength of the donor effect is a 46 percentage point difference in efficacy rate between high- and low-quality donors.

### Costello *et al*. 2019

In this study,^37^ 38 ulcerative colitis patients received FMT from 1 of 11 pools. Each pool included stool from 3 or 4 donors, out of 19 total donors. The outcome was steroid-free remission at 8 weeks. Similar to Paramsothy *et al*., there was no evidence of heterogeneity in patient outcomes by donor pool (*p* = 0.50, Fisher-Freeman-Halton test on 2 × 11 table) nor evidence of better outcomes for any particular donor (mid-*p* > 0.05 for all donors, Fisher’s exact test). Furthermore, a logistic regression estimated the difference between a very high quality pool (90th percentile in terms of efficacy) and low quality pool (10th percentile) as 0 percentage points but with a wide confidence interval (95% CI 0 to 74 percentage points).

### Jacob *et al*. 2017

In this study,^20^ 20 ulcerative colitis patients received FMT from 1 of 6 pools. Each pool included stool from 2 donors, out of 4 total donors. In this analysis, the outcome was clinical response at 4 weeks. Curiously, although a test for differences in patient outcomes by pool was statistically significant (Fisher-Freeman-Halton test on 2 × 6 table, *p* = 0.02), a test for differences in outcomes by donor was not (mid-p > 0.05 for all 4 donors, Fisher’s exact test). A logistic regression estimated the difference in efficacy between very high quality pools and low quality pools as very large (88 percentage points) but not statistically significant (95% confidence interval 0 to 100 percentage points).

There was no evidence for an association between patient outcomes and the bacterial community α-diversity of the pool they received (Figure 1a; *p* = 0.39, Mann-Whitney *U* test). In fact, more diverse pools were associated with worse patient outcomes. In our analysis, donor microbiota compositions also did not separate according to their associated patient outcomes (Figure 1b; *p* = 0.077, PERMANOVA), while they did in the original analysis (Figure 4C in that publication, *p* = 0.044), likely due to the differences in the 16S rRNA sequencing data processing pipeline. Our analysis used de-noising and the Bray-Curtis β-diversity metric, while Jacob *et al*. used 97% operational taxonomic units and the UniFrac metric.

**Figure 1:**
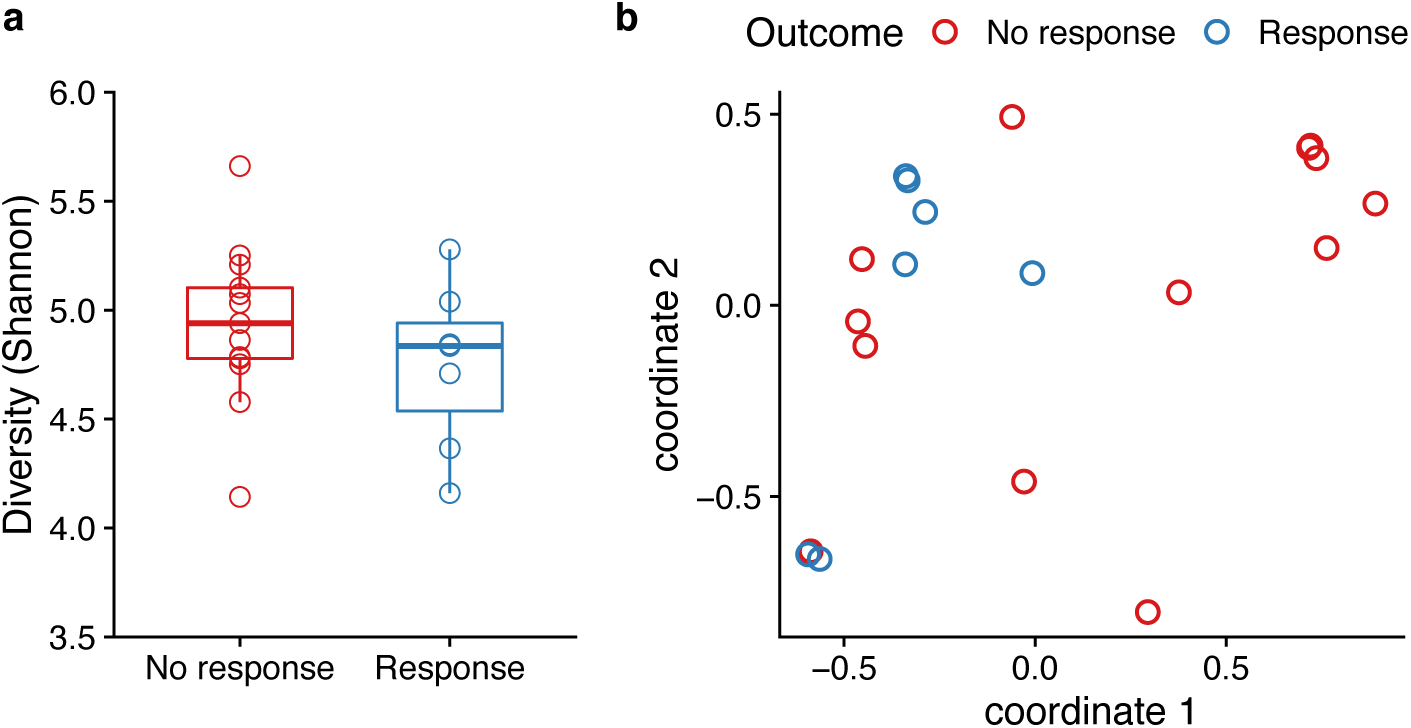
Microbiome analyses for Jacob *et al*. **a** Bacterial α-diversity of FMT pools do not significantly differ by patient outcomes (*p* = 0.39, Mann-Whitney *U* test). Each point represents a single patient. **b** Donor microbiota compositions (points; *x*-and *y*-coordinates show multidimensional scaling using Bray-Curtis dissimilarity) do not statistically significantly separate by associated patient outcome (color; *p* = 0.076, PERMANOVA). Each point represents the composition of the pool used to treat an individual patient.

### Meta-analysis of pool studies

Paramsothy *et al*., Costello *et al*., and Jacob *et al*. have sufficiently similar designs and the available data to allow a meta-analysis. A logistic regression on the combined data from all three studies, with the study as a fixed effect, estimated the difference in efficacy between very high quality pools and low quality pools as 41 percentage points but not statistically significant (95% CI 0 to 88 percentage points).

### Goyal *et al*. 2018

In this study,^24^ 21 patients with inflammatory bowel disease (any of Crohn’s disease, ulcerative colitis, or indeterminate colitis) received FMT and were available for follow-up. We used remission at 30 days as the outcome. Each patient received stool from a different donor, precluding any test of donor superiority. However, 16S rRNA sequencing was performed, allowing for an analysis of donation characteristic superiority, similar to Jacob *et al*. above. Similar to that study, there was no evidence for an association between patient outcomes and the α-diversity (Figure 2a; *p* = 0.97, Mann-Whitney *U* test) nor separation in donor microbiota (Figure 2b; *p* = 0.7, PERMANOVA).

**Figure 2.**
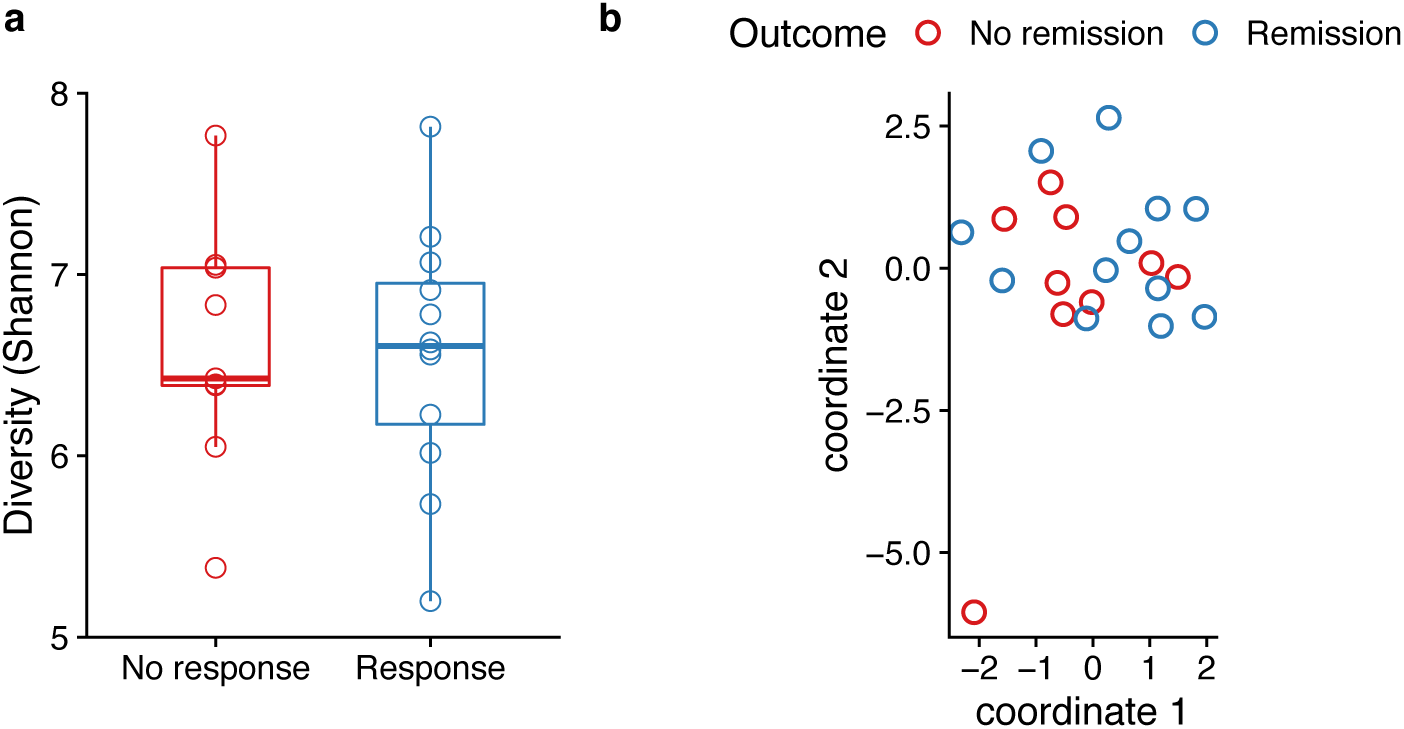
Microbiome analyses for Goyal *et al*. **a** Bacterial α-diversity of FMT pools do not significantly differ by patient outcomes (*p* = 0.97, Mann-Whitney *U* test). Each point represents a single patient. **b** Donor microbiota compositions (points; *x*-and *y*-coordinates show multidimensional scaling using Bray-Curtis dissimilarity) do not statistically significantly separate by associated patient outcome (color; *p* = 0.70, PERMANOVA). Each point represents the composition of the first sample from the donor used to treat an individual patient.

### Kump *et al*. 2017

In this study,^23^ 17 ulcerative colitis patients received 5 FMTs from 1 of 14 donors. 12 donors were used in 1 patient, 1 donor was used in 2 patients, and 1 donor was used in 3 patients. As in Goyal *et al*., this distribution of patients across donors does not permit an assessment of donor superiority. However, like Jacob *et al*. and Goyal *et al*., this study tested whether increased α-diversity was associated with improved patient outcomes.

We detected no difference in diversity by patient outcome (Figure 3a; *p* = 0.39, Kruskal-Wallis test; *p* > 0.05 for all 3 comparisons, Mann-Whitney *U* test), nor did we detect separation in donor microbiota according to associated patient outcomes (Figure 3b; *p* = 0.15, PERMANOVA).

**Figure 3:**
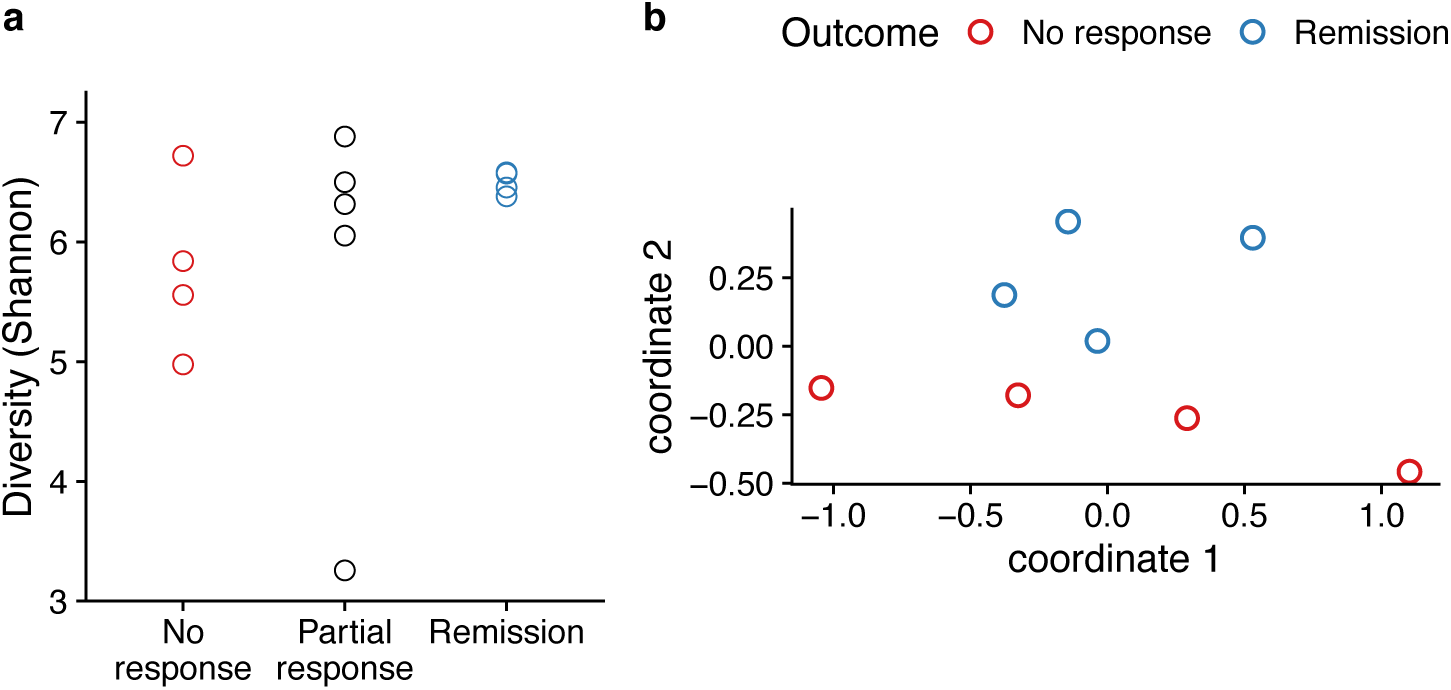
Microbiome analyses for Kump *et al*. **a** Bacterial α-diversity do not differ by patient outcome (*p* = 0.39, Kruskal-Wallis test; *p* > 0.05 for all 3 comparisons, Mann-Whitney *U* test). Each point represents a single patient. The diversity shown for each patient (vertical axis) is the average of the diversity of the sampled FMTs used in that patient. **b** Donor microbiota do not statistically significantly separate by associated patient outcome (color; *p* = 0.15, PERMANOVA). Each point represents the composition of the first donor sample administered to each patient.

The original publication reported statistically significant differences between the α-diversity of the 16 available donor stool samples associated with the 4 patients who achieved remission compared to the diversity of 12 available samples associated with the 4 patients who did not respond to FMT. Our analysis has a different conclusion, finding no support for a donor effect, principally because we analyzed the data as only 8 independent data points, that is, one data point for each patient (4 remission, 4 no response). By contrast, Kump *et al*. analyzed the data as if there were 28 independent data points, that is, one data point for each donation (12 associated with remission, 16 associated with no response). Using donations rather than patients to determine *N* is a statistical error known as “pseudoreplication”.^41^ Repeated measurements of the material delivered to a patient do not constitute independent measures of the patient’s outcome. In other words, when testing for an association between patient outcomes and a donation characteristic like α-diversity, the weight of the evidence is determined by the number of patient outcomes, not by the number of microbiome measurements.^42^

### Vermeire *et al*. 2016

In this study,^28^ 14 patients with inflammatory bowel disease (either of Crohn’s disease or ulcerative colitis) received FMT, each from a different donor. The original study reported that the bacterial α-diversity of donations varied by patient outcomes, that patients with successful outcomes received more bacterially diverse donations (Mann-Whitney *U* test, *p* = 0.012). However, the underlying bacterial sequencing data was not available for re-analysis.

### Nishida *et al*. 2017

In this study,^22^ 41 patients with ulcerative colitis received FMT, each from a different donor. The outcome was clinical response at 8 weeks. The original study found no difference in the α-diversity of the stool from 7 donors whose patients met the primary endpoint, compared to 19 donors whose patients did not (*p* = 0.69). The study also tested whether the abundance of 10 taxa were differentially abundant among those two groups of donors, finding *p*-values less than 0.05 for 2 of 9 taxa, neither of which is statistically significant after Benjamini-Hochberg multiple hypothesis correction. The underlying bacterial sequencing data was not available for re-analysis.

## DISCUSSION

In this study, we re-evaluated 9 studies that used FMT for IBD. In 7 of 9 cases, there was no statistically significant evidence of a donor effect. In one of the remaining studies (Jacob *et al*.), there was a statistically significant difference in efficacy by pool of donor stool, but not by donor, nor by the α-diversity of the stool or by donor microbiota composition. We were unable to re-analyze the other study (Vermeire *et al*.) that reported a statistically significant effect.

The major strengths of this study were the rigorous statistical approach used, which avoided pseudoreplication and misinterpretation of results from an adaptive trial design, and the uniformity of the re-analysis of the 16S rRNA microbiota composition data.

However, this study is also subject to two key limitations. First, our analysis considered only donor effects and made no reference to patient factors like disease history. Given the small sample sizes, adding patient factors as predictors would only decrease our ability to detect donor effects, which was contrary to our main aim. We do not mean to imply that all IBD patients are the same or should receive the same kind of treatment. In contrast, a principal conclusion from this study is that patient factors, not donor factors, are the main determinants of an FMT patient’s clinical outcome.

Second, our analysis considered only dichotomous patient outcomes and a small number of predictors, namely donors’ identities (e.g., donor A vs. donor B), pools’ identities (pool A vs. pool B), 16S rRNA microbiota composition α-diversity, and microbiota composition β-diversity. Previous work suggests that more proximal patient biomarkers, like mucosal immune markers, are more likely to be able to detect donor effects.^25^ We caution, however, that attempts to find associations between every conceivable donor factor, patient factor, and patient outcome risks “*p*-hacking”.^43^

Overall, we found only very weak evidence for donor effects. On the other hand, the small size of these studies —relative to the number of patients required to reliably detect a plausible donor effect^25^— means that the data cannot definitively rule out a large donor effect. It therefore remains undetermined if differences between donors are clinically relevant to FMT for IBD. We propose that larger studies, with careful biomarker selection and transparent reporting of results, is the best direction forward to characterizing the donor effect for FMT in IBD.

## Data Availability

Computer code and underlying data to reproduce the results are online at https://www.github.com/openbiome/donor-effects (DOI: 10.5281/zenodo.3780184).

## DECLARATIONS

## Acknowledgements

Sudarshan Paramsothy, Nadeem Kaakoush, and Rotem Sadovsky for assistance in collecting the Paramsothy *et al*. data and for helpful conversations. Sam Costello for assistance in collecting the Costello *et al*. data and for helpful conversations. Eric J. Alm, Shrish Budree, Claire Duvallet, Justin O’Sullivan, Pratik Panchal, Marina Santiago, Mark B. Smith, and Duane Wesemann for helpful conversations.

## Authorship statement

SWO is the guarantor. SWO conceived the study, obtained data, performed the analysis, and wrote the manuscript. YG obtained data; improved the analysis, interpretation, and presentation of the results; and revised the manuscript. All authors approved the final version of the manuscript.

## Statement of interests

SWO is employed by OpenBiome. YG is employed by and owns stock options in Finch Therapeutics. No specific funding was received for this work.

## Notes

### Competing Interest Statement

The authors have declared no competing interest.

### Funding Statement

SWO is an employee of OpenBiome. YG is an employee of Finch Therapeutics. No other funding was used for any aspect of the submitted work.

## REFERENCES

1. Marchesi, J. R. et al. The gut microbiota and host health: a new clinical frontier. Gut 65, 330–339 (2016).

2. Allegretti, J. R., Mullish, B. H., Kelly, C. & Fischer, M. The evolution of the use of faecal microbiota transplantation and emerging therapeutic indications. Lancet (London, England) 394, 420–431 (2019).

3. McDonald, L. C. et al. Clinical Practice Guidelines for Clostridium difficile Infection in Adults and Children: 2017 Update by the Infectious Diseases Society of America (IDSA) and Society for Healthcare Epidemiology of America (SHEA). Clin. Infect. Dis. 66, e1–e48 (2018).

4. Debast, S. B., Bauer, M. P. & Kuijper, E. J. European Society of Clinical Microbiology and Infectious Diseases: Update of the Treatment Guidance Document for Clostridium difficile Infection. Clin. Microbiol. Infect. 20, 1–26 (2014).

5. Olesen, S. W., Panchal, P., Chen, J., Budree, S. & Osman, M. Global disparities in faecal microbiota transplantation research. Lancet Gastroenterol. Hepatol. 5, 241 (2020).

6. Lopetuso, L. R. et al. Fecal transplantation for ulcerative colitis: current evidence and future applications. Expert Opin. Biol. Ther. 20, 343–351 (2020).

7. David, L. A. et al. Diet rapidly and reproducibly alters the human gut microbiome. Nature 505, 559–563 (2014).

8. Turnbaugh, P. J. et al. The human microbiome project. Nature 449, 804–10 (2007).

9. Rossen, N. G. et al. Findings From a Randomized Controlled Trial of Fecal Transplantation for Patients With Ulcerative Colitis. Gastroenterology 149, 110–118.e4 (2015).

10. Wilson, B. C., Vatanen, T., Cutfield, W. S. & O’Sullivan, J. M. The Super-Donor Phenomenon in Fecal Microbiota Transplantation. Front. Cell. Infect. Microbiol. (2019) doi:10.3389/fcimb.2019.00002.

11. Olesen, S. W., Leier, M. M., Alm, E. J. & Kahn, S. A. Searching for superstool: Maximizing the therapeutic potential of FMT. Nature Reviews Gastroenterology and Hepatology (2018) doi:10.1038/s41575-018-0019-4.

12. Olesen, S. W., Gurry, T. & Alm, E. J. Designing fecal microbiota transplant trials that account for differences in donor stool efficacy. Stat. Methods Med. Res. (2018) doi:10.1177/0962280216688502.

13. Duvallet, C. et al. Framework for rational donor selection in fecal microbiota transplant clinical trials. PLoS One 14, e0222881 (2019).

14. Barnes, D. et al. Competitively Selected Donor Fecal Microbiota Transplantation. J. Pediatr. Gastroenterol. Nutr. 67, 185–187 (2018).

15. Osman, M., Abend, A., Panchal, P., Kassam, Z. & Budree, S. Does the Donor Matter? Microbiome Sequencing to Evaluate Lower Donor Efficacy in Fecal Microbiota Transplantation for Recurrent Clostridium Difficile Infection. Gastroenterology 154, S-25-S-26 (2018).

16. Ray, A. & Jones, C. Does the donor matter? Donor vs patient effects in the outcome of a next-generation microbiota-based drug trial for recurrent Clostridium difficile infection. Future Microbiol. 11, 611–616 (2016).

17. Zuo, T. et al. Gut fungal dysbiosis correlates with reduced efficacy of fecal microbiota transplantation in Clostridium difficile infection. Nat. Commun. 9, 3663 (2018).

18. Moayyedi, P. et al. Fecal Microbiota Transplantation Induces Remission in Patients With Active Ulcerative Colitis in a Randomized Controlled Trial. Gastroenterology 149, 102–109.e6 (2015).

19. Paramsothy, S. et al. Multidonor intensive faecal microbiota transplantation for active ulcerative colitis: a randomised placebo-controlled trial. Lancet 389, 1218–1228 (2017).

20. Jacob, V. et al. Single Delivery of High-Diversity Fecal Microbiota Preparation by Colonoscopy Is Safe and Effective in Increasing Microbial Diversity in Active Ulcerative Colitis. Inflamm. Bowel Dis. 23, 903–911 (2017).

21. Paramsothy, S. et al. Faecal Microbiota Transplantation for Inflammatory Bowel Disease: A Systematic Review and Meta-analysis. J. Crohn’s Colitis 11, 1180–1199 (2017).

22. Nishida, A. et al. Efficacy and safety of single fecal microbiota transplantation for Japanese patients with mild to moderately active ulcerative colitis. J. Gastroenterol. 52, 476–482 (2017).

23. Kump, P. et al. The taxonomic composition of the donor intestinal microbiota is a major factor influencing the efficacy of faecal microbiota transplantation in therapy refractory ulcerative colitis. Aliment. Pharmacol. Ther. 47, 67–77 (2018).

24. Goyal, A. et al. Safety, Clinical Response, and Microbiome Findings Following Fecal Microbiota Transplant in Children With Inflammatory Bowel Disease. Inflamm. Bowel Dis. 24, 410–421 (2018).

25. Olesen, S. W. Power calculations for detecting differences in efficacy of fecal microbiota donors. medRxiv (2020) doi:10.1101/2020.04.16.20068361.

26. Wasserstein, R. L., Schirm, A. L. & Lazar, N. A. Moving to a World Beyond “ p < 0.05”. Am. Stat. 73, 1–19 (2019).

27. Budree, S. et al. The Association of Stool Donor Diet on Microbial Profile and Clinical Outcomes of Fecal Microbiota Transplantation in Clostridium Difficile Infection. Gastroenterology 152, S630–S631 (2017).

28. Vermeire, S. et al. Donor Species Richness Determines Faecal Microbiota Transplantation Success in Inflammatory Bowel Disease. J. Crohn’s Colitis 10, 387–394 (2016).

29. Kazerouni, A. & Wein, L. M. Exploring the Efficacy of Pooled Stools in Fecal Microbiota Transplantation for Microbiota-Associated Chronic Diseases. PLoS One 12, e0163956 (2017).

30. Khanna, S. et al. Changes in microbial ecology after fecal microbiota transplantation for recurrent C. difficile infection affected by underlying inflammatory bowel disease. Microbiome 5, 55 (2017).

31. R Core Team. R: A language and environment for statistical computing. (R Foundation for Statistical Computing, 2019).

32. Fay, M. P. Confidence intervals that match Fisher’s exact or Blaker’s exact tests. Biostatistics 11, 373–374 (2010).

33. Mehta, C. R. & Hilton, J. F. Exact power of conditional and unconditional tests: Going beyond the 2×2 contingency table. Am. Stat. (1993) doi:10.1080/00031305.1993.10475946.

34. Anderson, M. J. A new method for non-parametric multivariate analysis of variance. Austral Ecol. 26, 32–46 (2001).

35. Oksanen, J. et al. vegan: Community Ecology Package. https://cran.r-project.org/package=vegan (2019).

36. Bates, D., Mächler, M., Bolker, B. M. & Walker, S. C. Fitting linear mixed-effects models using lme4. J. Stat. Softw. (2015) doi:10.18637/jss.v067.i01.

37. Costello, S. P. et al. Effect of Fecal Microbiota Transplantation on 8-Week Remission in Patients With Ulcerative Colitis. JAMA 321, 156 (2019).

38. Bolyen, E. et al. Reproducible, interactive, scalable and extensible microbiome data science using QIIME 2. Nat. Biotechnol. 37, 852–857 (2019).

39. Amir, A. et al. Deblur Rapidly Resolves Single-Nucleotide Community Sequence Patterns. mSystems 2, (2017).

40. Bhatt, D. L. & Mehta, C. Adaptive Designs for Clinical Trials. N. Engl. J. Med. 375, 65–74 (2016).

41. Hurlbert, S. H. Pseudoreplication and the Design of Ecological Field Experiments. Ecol. Monogr. 54, 187–211 (1984).

42. Lazic, S. E., Clarke-Williams, C. J. & Munafò, M. R. What exactly is ‘N’ in cell culture and animal experiments? PLOS Biol. 16, e2005282 (2018).

43. Head, M. L., Holman, L., Lanfear, R., Kahn, A. T. & Jennions, M. D. The Extent and Consequences of P-Hacking in Science. PLOS Biol. 13, e1002106 (2015).

